# Exploring county-level spatio-temporal patterns in opioid overdose related emergency department visits

**DOI:** 10.1101/2022.05.24.22275495

**Authors:** Angeela Acharya, Alyssa M Izquierdo, Stefanie F Gonçalves, Rebecca A Bates, Faye S Taxman, Martin P Slawski, Huzefa S Rangwala, Siddhartha Sikdar

## Abstract

Opioid overdoses within the United States continue to rise and have been negatively impacting the social and economic status of the country. In order to effectively allocate resources and identify policy solutions to reduce the number of overdoses, it is important to understand the geographical differences in opioid overdose rates and their causes. In this study, we utilized data on emergency department opioid overdose (EDOOD) visits to explore the county-level spatio-temporal distribution of opioid overdose rates within the state of Virginia and their association with aggregate socio-ecological factors. The analyses were performed using a combination of techniques including Moran’s I and multilevel modeling. Using data from 2016-2021, we found that Virginia counties had notable differences in their EDOOD visit rates with significant neighborhood-level associations: many counties in the southwestern region were consistently identified as the hotspots (areas with a higher concentration of EDOOD visits) whereas many counties in the northern region were consistently identified as the coldspots (areas with a lower concentration of EDOOD visits). In most Virginia counties, EDOOD visit rates declined from 2017 to 2018. In more recent years (since 2019), the visit rates showed an increasing trend. The multilevel modeling revealed that the change in clinical care factors (i.e., access to care and quality of care) and socio-economic factors (i.e., levels of education, employment, income, family and social support, and community safety) were significantly associated with the change in the EDOOD visit rates. The findings from this study have the potential to assist policymakers in proper resource planning thereby improving health outcomes.

## Introduction

The continued rise in drug overdoses involving opioids has significantly impacted the social and economic fabric of communities in the United States [1]. For instance, in 2017, the economic cost of opioid deaths, criminal justice involvement, treatment, and lost wages surpassed $1.02 trillion [2]. Between April 2020 and April 2021, over 100,000 deaths involved opioids--an increase of 28.5% over the previous 12 months [3]. An effective response to the opioid crisis requires policymakers to understand which communities are the most affected and how the resources have been allocated in those communities [4].

Recent research has identified many socio-ecological factors as the risk factors of opioid overdose [5-6]. These risk factors are inextricably linked to personal and environmental factors and systems that hinder rather than support individuals [6]. For instance, among individuals receiving healthcare services at a free clinic, prescription opioid misuse was more likely among patients who were employed and less likely among those with post-high school education [7]. Uninsured individuals were significantly more likely than insured individuals to be high-risk drug users [8]. Access to prescription opioids is another prominent risk factor of unintentional opioid overdose deaths [9-10], which has increased in rural areas with a greater need for medical services [11-12].

Opioid overdose rates within the United States have been found to vary across different geographical regions. A socio-ecological framework posits that the characteristics of the community in which individuals live significantly influence their health behaviors [13]. For instance, counties across the United States with high economic distress, high opioid prescription rates, and a lack of opioid treatment program providers have higher opioid overdose mortality rates [14-15]. Statewide health disparities, including lower socioeconomic status and access to health care, can differ at the county level and are more predominant in rural areas [16]. Opioid overdose patterns are consistent with counties that may lack the resources necessary to prevent overdose [17]. In sum, the intersection of where and how people live is a significant factor in health outcomes and requires public health to work with urban planning to create supportive and healthy environments to reduce the risks of opioid overdose [18].

Besides geographical differences, there have been significant variations in the temporal trends of opioid overdose rates. Beginning around the year 2000, opioid overdose rates in the United States have been increasing over time [19]. In recent years (since the early 2010s), much of this growth has been attributed to synthetic opioids such as fentanyl, which increased more than 50% from 2019 to 2020 [20]. During the same time period, prescription opioid-related overdose showed the first increase in years (10.6%) whereas heroin-related overdose showed a downward trend (down 3.6%), similar to the recent prior years [20]. Not all regions within the United States have the same characteristics and the national pattern may not well reflect the local growth trajectories.

Prior research has implemented different spatiotemporal analysis techniques to identify the local geographical differences in opioid overdose rates over time. For instance, Hernandez et al. [21] examined prescription opioid death rates in Ohio from 2010-2017 and identified 12 hotspots along with three significant changing trends of opioid overdose using temporal trend analysis. Marotta et al. [17] examined cumulative opioid overdose deaths in New York State using data from 2013-2015 and identified geographical hotspots of overdose death rates for different types of opioids. Sauer et.al [22] used spatiotemporal bayesian modeling and exploratory spatial analysis to evaluate risk factors related to drug-involved emergency department visits in the greater Baltimore metropolitan from 2016-2019. These studies and a few others [23-25] have demonstrated that spatio-temporal techniques utilizing local population-level data can provide a profile of opioid overdose risk.

In this study, we performed a county-level spatio-temporal assessment of opioid overdose rates and their association with different socio-ecological factors for the state of Virginia. We focus on Virginia for two main reasons. First, its growing rates of fatal opioid overdose [26] represent the growing rates of opioid overdose across the United States. Secondly, the Virginia Department of Health collects monthly emergency department opioid overdose (EDOOD) visit data as part of syndromic surveillance to measure health trends [27]. This publicly available dataset can serve as a timely indicator of opioid overdose trends. Different from prior studies, we used the EDOOD visits as a proxy for total overdoses to understand the spatio-temporal dynamics of opioid overdose rates and potential socio-ecological risk and protective factors. Emergency departments (EDs) are the primary treatment venue for patients with overdoses [28]. In recent years, urgent care centers’ utilization to treat opioid overdoses has also significantly increased. Between 2007-2016 documented claims for urgent care centers increased by 1,725 percent compared to a 229 percent increase for emergency department claims with ‘injury, poisoning, and consequences of external causes’ [29]. However, the current study only focuses on the visits obtained from hospital-based and free-standing EDs. EDOOD visit rates increased by 28.5 percent across the United States in 2020, compared to 2018 and 2019 [30]. Understanding the spatio-temporal trends of EDOOD visit rates can help identify targets for policy change and timely resource allocation to mitigate the opioid crisis.

To summarize, this study utilized a comprehensive, three-pronged approach to understanding the opioid overdose trends in Virginia. The goals of the study were to 1) identify spatio-temporal variations of EDOOD visit rates from 2016-2021 among Virginia counties, 2) assess how counties cluster together based on their EDOOD visit rates, and 3) identify socio-ecological factors that are associated with the change in EDOOD visit rates over time. Moran’s I [31], Local Indicators of Spatial Association (LISA) [32], Dynamic Time Warping (DTW) [33], and multilevel modeling [34] were implemented for the spatio-temporal analysis. To our knowledge, this is the first study to combine techniques from statistics, data mining, and geographic information systems (GIS) to examine how a county performs in terms of EDOOD visits. Although the study focuses on Virginia, the study methods can be extended to other geographic locations with similar data. The source code can be made available upon request.

## Methods and Materials

### Study area

We analyzed the EDOOD visit rates and the associated socio-ecological factors across the different counties within the state of Virginia, United States. The state of Virginia consists of 95 counties and 38 independent cities that are considered county-equivalent for census purposes. The analysis was performed for those 133 unique geographic regions.

### Measures

#### Emergency department opioid overdose visits

The EDOOD visits dataset was obtained from the Virginia Department of Health (VDH) [35] and is based on syndromic surveillance reported by hospitals and free-standing EDs in Virginia. It consists of count and rate statistics (monthly and annual) of ED visits for unintentional opioid overdose (fatal and nonfatal) among Virginia residents aggregated by different geographical units. This dataset separates heroin from all other types of opioid overdoses. In Virginia, only a small percentage of the total overdoses are driven by heroin in recent years [35]. The numbers are not significant enough to analyze the spatio-temporal variations of opioid overdose rates. Thus, this study did not include heroin-related EDOOD visits in its analyses.

Although the EDOOD data spans from 2015 to 2022, we only examined data from 2016 to 2021 given that complete data (i.e., previous 12-month average visits, annual summary) was not available for 2015 and 2022. The outcome variable was defined as the rate of EDOOD visits per 100,000 population.

#### Geography assignment

The geographic location in the EDOOD visit dataset is assigned based on the patient’s self-reported residential zip code. A single zip code may belong to multiple cities and counties. In that case, the visit is assigned to the locality where the majority of the population resides. Additionally, some Virginia cities and counties are aggregated (e.g., Alleghany County and Covington City) due to overlapping zip codes (see Appendix).

#### EDOOD visit definition

ICD9 (e.g., 965.00, 965.01), ICD10 (e.g., T40.0×1A, T40.0×4A), or SNOMED (e.g., 295165009, 242253008) codes representing opioid overdose were used to identify overdose incidents. Similarly, the mention of terms like Narcan or naloxone in the chief complaint or discharge diagnosis was also used to identify overdoses. This includes unintentional overdose by opioids or unspecified substances (excluding heroin). For complete information on the case definition, please refer to [36].

#### Converting visit counts to rates

Whenever information was only available in the form of counts, we used the population data from American Community Survey (ACS) to calculate the rates (visits per 100K population) [37]. We followed the same guidelines as specified in [35] for the population rates: 2016-2017 rates are based on the corresponding ACS year estimates, 2018 and 2019 rates are based on 2017 estimates, 2020 rates are based on 2018 estimates, and 2021 rates are based on 2019 estimates.

#### Socio-ecological factors

Socio-ecological factors at the Virginia county level were obtained from the County Health Rankings & Roadmaps (CHR&R) dataset from the Robert Wood Johnson Foundation [38]. We extracted data from 2016 to 2021 to be consistent with the EDOOD visit dataset. This publicly available dataset aggregates data from the Centers for Disease Control and Prevention (CDC) as well as other sources (e.g., U.S. Census Bureau, Behavioral Risk Factor Surveillance System) to provide yearly county-level rankings based on different attributes related to health outcomes. Based on prior studies on the association between socio-ecological factors and opioid overdoses [5–9, 14-15], we selected four variables: health behaviors, clinical care, social and economic factors, and physical environment as the most appropriate for our study (see Fig 1). The goal was to examine if these socio-ecological factors influence the spatiotemporal trends of EDOOD visit rates. Other sources have also aggregated data on socio-ecological factors (e.g., Opioid Environment Policy Scan, Virginia Department of Health, etc.). However, the CHR&R dataset is an adequate proxy for socio-ecological factors for our analysis because it encompasses a wide range of socio-ecological inputs within its four variables:

**Fig 1.**
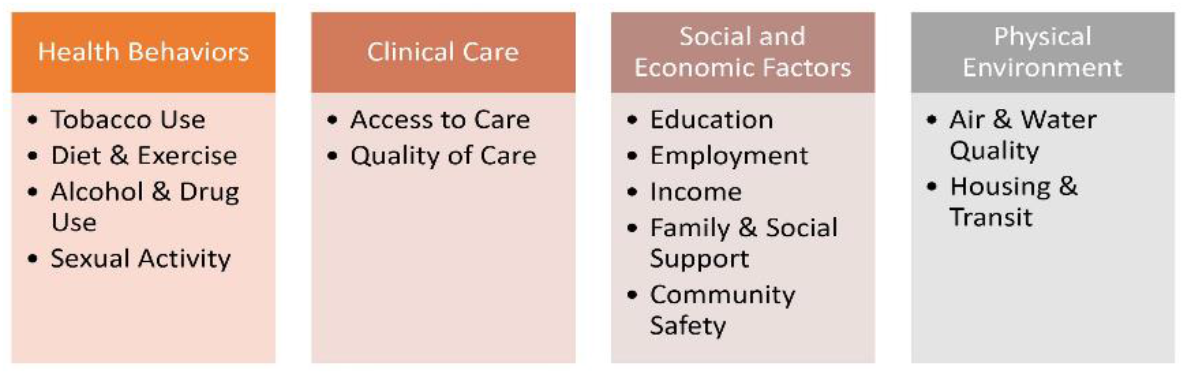
Socio-ecological factors from County Health Rankings & Roadmaps (CHR&R)

##### Clinical care

Includes inputs about access to care and quality of care, such as uninsured rates, and access to primary care physicians, dentists, and mental health providers.

##### Social and economic factors

Includes inputs from six unique data sources measuring unemployment, children in poverty, income inequality, single-parent households, social associations, violent crime, and injury deaths.

##### Physical environment

Includes inputs from four unique data sets measuring air pollution, alcohol drinking violations, severe housing problems, driving alone to work, and long commute-driving alone.

##### Health behaviors

Includes inputs about tobacco and alcohol use, diet and exercise, and sexual activity from seven unique data sources. These inputs further include physical inactivity, excessive alcohol use and impaired driving deaths, sexually transmitted diseases, and teen pregnancy.

The rankings in the CHR&R dataset were calculated using standardized z-scores from several data sources [39]. The county with the lowest z-score received a rank of 1, which indicates the highest quality of socio-ecological factors (e.g., low tobacco use, better access to care, better education). The county with the highest z-score received a ranking of 133, which indicates the lowest quality of socio-ecological factors (e.g., high tobacco use, poor access to care, poor education). The z-scores were averaged for the counties (or cities) that were combined in the EDOOD visit dataset. The CHR&R dataset has been validated in other studies [40-41].

##### Neighborhood adjacency

Data on neighborhood adjacency for Virginia counties was obtained from the US Census Bureau [42]. This data lists each county along with its adjoining neighbors including counties that are not in Virginia but adjacent to Virginia counties. For this study, we only considered the neighboring counties that are a part of Virginia. This data was utilized for our spatial analysis.

## Data Analysis

### Spatial analysis

To identify any spatial variations in the opioid overdose rates across Virginia counties, we calculated the spatial autocorrelation using data on neighborhood adjacency and EDOOD visits for the years 2016-2021. Average monthly EDOOD visit rates were used to summarize the yearly EDOOD trends. Spatial autocorrelation is the phenomenon where the presence of some quantity in an area makes its presence in neighboring areas more or less likely [43]. Positive autocorrelation, which is more common in practice, is the tendency for areas that are close together to have similar values. In contrast, negative autocorrelation is the tendency for areas that are close together to have different values.

### Global spatial autocorrelation

Global auto-correlation measures the overall association within the data. In this study, it measures the similarity between the neighboring counties in terms of EDOOD visit rates. We calculated the Moran’s I index, a common measure of global auto-correlation [31]. Then we performed a permutation test to assess the significance of Moran’s I index analysis. The values of Moran’s I range from +1 (strong positive spatial auto-correlation) to 0 (randomness) to -1 (strong negative pattern). Moran’s I value of 0.7, for instance, indicates that the spatial pattern across counties is homogeneous meaning that the neighboring counties have very similar visit rates.

### Local spatial autocorrelation

To study the contribution of each county to the global Moran’s I index and identify local hotspots (clusters of high EDOOD visit rates) and coldspots (clusters of low EDOOD visit rates), we calculated Local Indicators of Spatial Association (LISA) [32] for each county. These auto-correlation indices were used to divide the counties into four distinct groups:

- High-high: *Counties* with high visit rates with *neighboring counties* that also have high visit rates (also known as hotspots)
- Low-low: *Counties* with low visit rates with *neighboring counties* that also have low visit rates (also known as coldspots)
- High-low: *Counties* with high visit rates but surrounded by *counties* that have low visit rates
- Low-high: *Counties* with low visit rates but surrounded by *counties* with high visit rates

The classification of counties into regions of low or high visit rates was done based on whether they had rates less than or greater than (or equal to) the mean value of visit rates across the state of Virginia. A permutation test was used to identify non-significant associations within neighbors (counties) thereby highlighting the significant hotspots and coldspots of EDOOD visit rates. Both local and global autocorrelation analyses were performed in python using the PySAL package [44].

### Temporal analysis

Temporal analysis refers to the study of an outcome over time. We used two different methods-- clustering and multi-level modeling--to analyze the EDOOD visit rates over time and identify their association with different socio-ecological risk factors.

### Clustering

To identify similarities between the temporal trends of visit rates in different counties, we used dynamic time warping (DTW) [33]. DTW is a data mining technique used to compute the similarity between multiple time series (e.g., opioid overdose rates over time) and cluster them together based on their shapes and magnitudes. We utilized the moving average of the monthly visit rates (for a smoother curve) to perform the clustering. After we obtained clusters of counties, we mapped the clusters using choropleth maps for easy visualization. Analyses were performed in Python using the PySAL package and tslearn package [45].

### Multilevel modeling

To identify how the changes in EDOOD visit rates relate to the changes in different socio-ecological factors, we performed multilevel modeling [34] with the visit rates as the outcome variable and four different time-varying aggregated variables from the CHR&R dataset as our predictors. Multilevel modeling is advantageous because it accounts for correlations across time within individual counties. Moreover, multilevel models handle missing data in visit rates for any county and at any time point without pairwise deletion of individual counties. Multilevel modeling was performed in R using the lme4 package [46]. A forward-stepping procedure was used to create the final model [47]. First, an unconditional means model (i.e., baseline model) was created. From this model, an intraclass correlation coefficient (ICC)--representing the proportion of variance explained within counties--was computed. Next, conditional growth models were created to examine the linear effect of time on EDOOD visit rates, with time modeled as a fixed and random slope in separate models. The model (i.e., either fixed or random slope) with a better fit compared to the unconditional growth model was used moving forward. Finally, a conditional random growth model was created to examine the linear effects of the time-varying covariates (i.e., socio-ecological factors) on the visit rates:

Level 1:

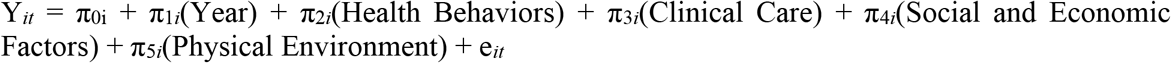

Level 2:

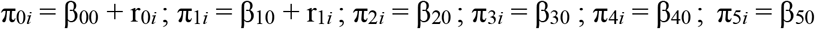

The Level-1 equation models the within-county variance based on EDOOD visit rates. Thus, for county *i* at time *t*, the expected outcome, Y, is equal to the intercept, π_0*i*_, plus an effect for the slope, π_1*i*_, plus an effect for Health Behaviors, π_2*i*_, plus an effect for Clinical Care, π_3*i*_, plus an effect for Social and Economic Factors, π_4*i*_, plus an effect for Physical Environment, π_5*i*_, plus error, e_*it*_. The Level-2 equations state that the intercept and year were fitted using random effects whereas the socio-ecological predictors were modeled using fixed effects. We used a fixed slope for the socio-ecological factors because specifying many random coefficients overfits the model producing misleading results [48]. Lastly, we ran ANOVA-like table with tests of random effects for each model using the ImerTest package. Each model was compared to the preceding model using these ANOVA tests.

## Results

### Spatial analysis

#### Global auto-correlation

The results from Moran’s I index indicate that there is some similarity between counties and their neighbors with respect to their EDOOD visit rates in most of the years. As shown in Table 1, the values of indices are greater than 0 for all the years indicating a positive global spatial autocorrelation. However, the similarity scores and the corresponding p-values vary across different years. The strongest association seems to be present in the year 2018 (Moran’s I = 0.25). This indicates that many neighboring counties had similar EDOOD visit rates during that time. On the other hand, the Morans’I value for 2016 is almost close to 0, meaning that the EDOOD visit rates were not significantly similar across the neighboring counties.

**Table 1.** Global moran’s I indices returned for the years 2016-2019.

| Year | Moran's I Index | p-value |
| --- | --- | --- |
| 2016 | 0.04 | 0.182 |
| 2017 | 0.11 | 0.033 |
| 2018 | 0.25 | 0.001 |
| 2019 | 0.07 | 0.089 |
| 2020 | 0.09 | 0.04 |
| 2021 | 0.10 | 0.04 |

*Note.* Positive value: Neighboring counties have homogeneous patterns. Negative value: Neighboring counties have heterogeneous patterns.

#### Local auto-correlation

Fig 2 represents the plots of EDOOD visit rates alongside the classification returned by LISA for the 3 years (2017, 2018, and 2021) with the most significant neighborhood association (based on Moran’s I values). To show them in the maps, we assigned the same value/color coding to counties that were combined together in the analyses (although they were only counted once in analyses). For instance, the combined rate for Grayson and Galax for the year 2016 was 21.7, but they are both represented separately as a value of 21.7 on the map.

**Fig 2.**
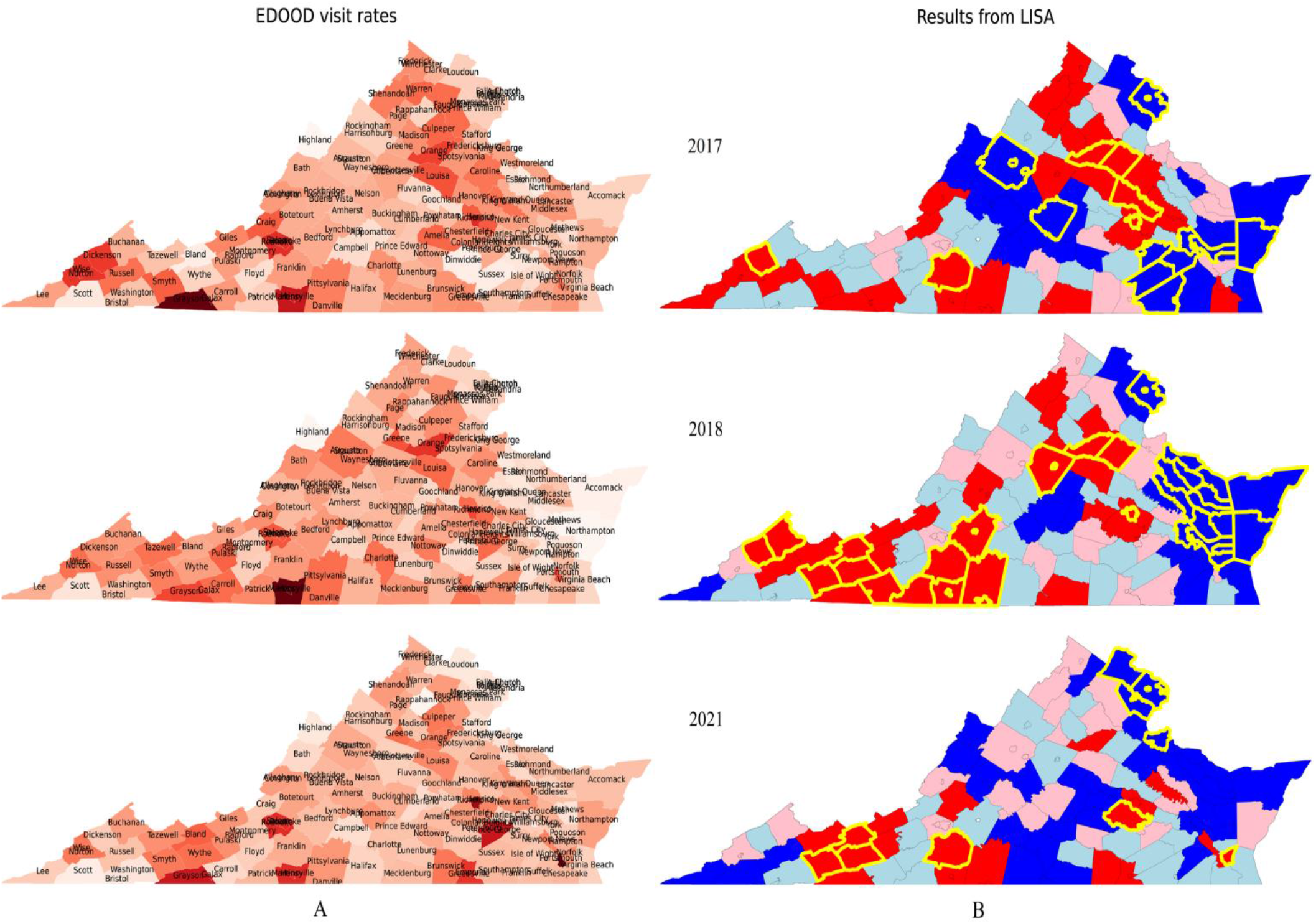
Choropleth plots of (A) EDOOD visit rates (darker color signifies higher rates), and (B) results from LISA analysis returned for the years 2016-2021 highlighted by different colors. (red: high-high – hot spots, blue: low-low – cold spots, pink: high-low, light blue: low-high). The yellow border marks the significant clusters.

#### EDOOD visit rates

As shown in Fig 2A, The EDOOD visit rates differed within the state and across time. However, some of the highest rates of visits can be seen across the southwestern region (e.g., Galax & Grayson, Smyth, Martinsville & Henry) and also in the northwestern region (e.g., Orange, Louisa, Culpeper). In contrast, most counties in the northern region (e.g., Fairfax & Falls Church, Loudon) and some in the eastern region (e.g., Accomack) had relatively lower rates.

#### LISA values

The LISA values were calculated for every county in Virginia for the years 2016-2021. However, as aforementioned, Fig. 2B only presents results for the years 2017, 2018, and 2021. The four distinct subgroups returned by LISA are shown in choropleth maps (in Fig. 2B) in distinct colors: red (high-high), blue (low-low), light blue (low-high), pink (high-low). The significant clusters returned by the permutation test are highlighted in yellow. These highlighted counties had significantly higher or lower concentrations of EDOOD visit rates based on their color coding (red: hotspots, blue: coldspots). As expected, the hotspots were mostly concentrated around the southwestern region and the northwestern region. The cold spots were scattered across multiple regions with the most consistent one being in the northern region. There were some counties (pink clusters and blue clusters) that had significantly different visit rates than their neighboring counties. For instance, counties like Floyd, Scott, and Caroll in southwestern Virginia had lower EDOOD visit rates even when most of their neighboring counties had higher rates. Similarly, as also verified by Moran’s I, the neighborhood similarity seems to be the most prominent in the year 2018, where there are close clusters of high and low EDOOD visit rates. The locations of the LISA subgroups seem to be changing over time.

### Temporal analysis

#### Clustering

Five distinct groups of counties (clusters) were returned by the DTW clustering algorithm. The clusters with plots of their temporal trends and their corresponding geospatial mapping are provided in Fig 3. These clusters differ in their magnitude and trend over time. As depicted in Fig. 3, counties in Groups A and E had similar trends over time but differed in their magnitudes. Most counties in these groups had EDOOD visit rates that slightly decreased from 2016 to 2018, which started increasing around 2019. However, counties in Group A (e.g., Buchanan, Louisa, Shenandoah) had slightly lower rates of EDOOD visits (starting range 7-16) as compared to the counties in Group E (e.g., Orange, Smyth, Wise, Dickenson), which had a starting range of 13-20. Similarly, counties in Group B (e.g., Amelia, Bland, Amherst) and Group C (e.g., Grayson & Galax, Martinsville & Henry) also had similar trends which differed in magnitudes. Overall, these counties had increasing rates over time. Group B had a starting range of 0-10 whereas Group C had a starting range of 0-15. Group C, however, had a steeper rising curve with an ending range of 16-38. Lastly, counties in Group D (e.g., Arlington, Fairfax & Falls Church, Alexandria, Williamsburg) did not show a consistent temporal trend and mostly had low EDOOD visit rates throughout the 6-year period. Note that for each month, we plotted the average EDOOD visit rates of the previous 12 months.

**Fig 3.**
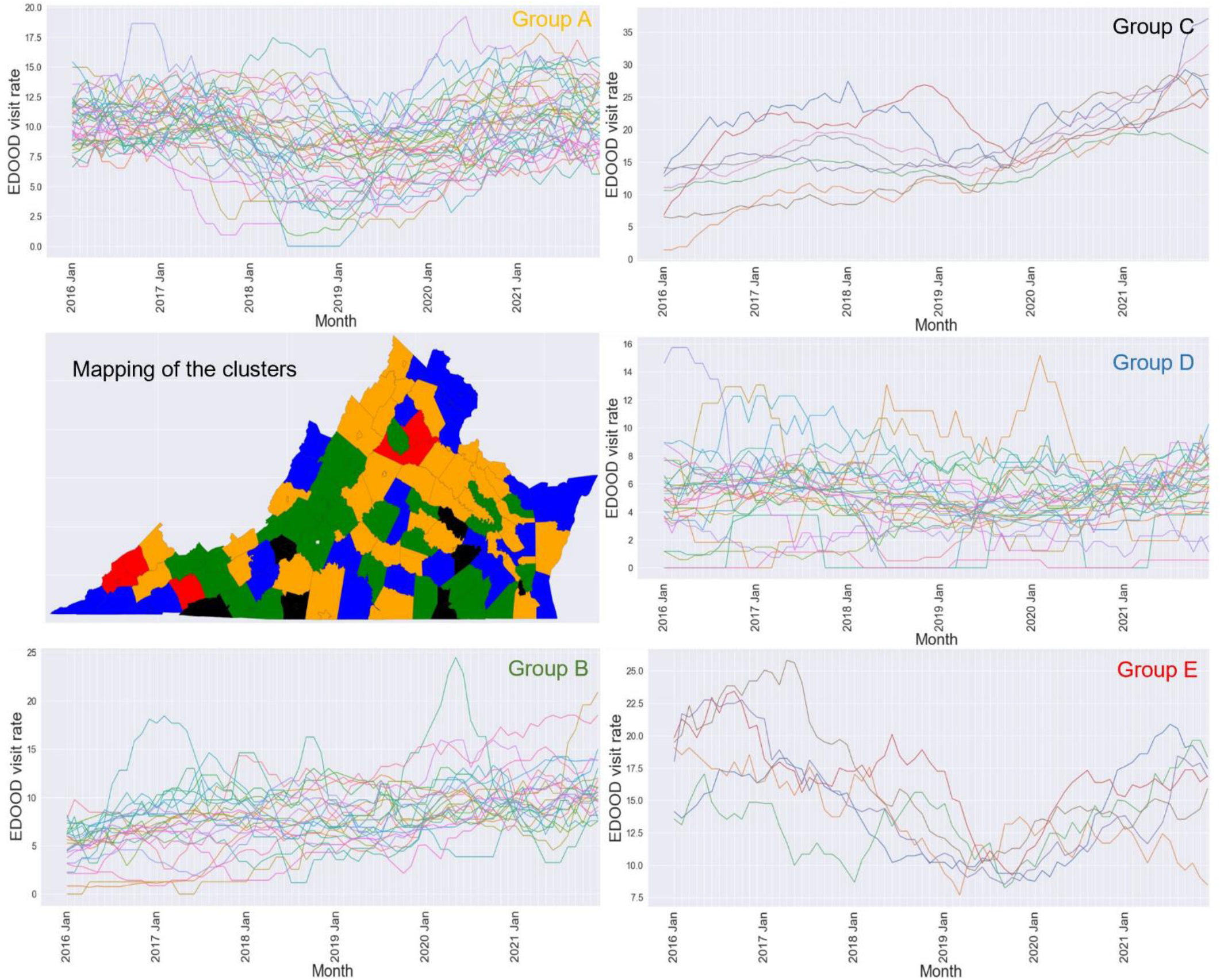
5 different cluster groups (A, B, C, D, E) as determined by the Dynamic Time Warping (DTW) clustering algorithm along with their corresponding choropleth mapping. *Note*. Each cluster group is represented by a unique color in the choropleth map. The same color is used to represent the cluster group names.

### Multilevel modeling

The baseline unconditional model returned an ICC of 0.59, which indicates that 59% of the variance is attributable to differences between counties while 41% of the variance is attributable to differences within counties over time. Given that more than 5% of the variance is attributable to differences within counties over time, the use of multilevel modeling is justified. We ran ANOVA-like table tests of random effects for each model and compared each model to the preceding model. First, we found that the fixed growth model was a better fit than the unconditional model (*X*² = 34.37, df = 1, Pr(>Chisq) = 4.556 × 10^−9^, *p* < .001). Next, we found that the random growth model was a better fit than the fixed growth model (*X*² = 60.197, *df* = 2, Pr(>Chisq) = 8.481 × 10^−14^, *p* < 0.001). Thus, we proceeded to run a random growth model with our time-varying socio-ecological predictors. Table 2 shows the goodness of fit values for all four models. The random growth model showed that time predicted, on average, increases in EDOOD visits from 2016-2021 (see Table 2). When incorporating predictors into the model (i.e., conditional random growth model), Clinical Care and Social and Economic Factors emerged as significant time-varying predictors of the slope for EDOOD visits (see Table 2). This suggests that a 1 unit decrease in Clinical Care z-scores (i.e., higher ranking/better clinical care) increased the slope of EDOOD visit rates by 6.64. In addition, a 1 unit increase in Social and Economic Factors z-scores (i.e., lower ranking/lower quality of social and economic factors) increased the slope of visit rates by 6.74. These sociological factors are important predictors of changes in EDOOD visits.

**Table 2.** Multilevel growth models with socio-ecological risk factors as time-varying covariates (TVCs)

| Parameters | Unconditional | Fixed Growth | Random Growth | Random Growth with TVCs |
| --- | --- | --- | --- | --- |
| *Fixed Effects* | $\beta$ (SE) | $\beta$ (SE) | $\beta$ (SE) | $\beta$ (SE) |
| Intercept | 9.14(0.39)*** | 8.08(0.42)*** | 8.08(0.38)*** | 8.14(0.37)*** |
| Time (Year) | — | 0.42(0.07)*** | 0.42(0.10)** | 0.42(0.10)*** |
| Health Behaviors | — | — | — | -3.36(2.41) |
| **Clinical Care** | — | — | — | **-6.64(3.04)\*** |
| **Social and Economic Factors** | — | — | — | **6.74(1.83)\*\*** |
| Physical Environment | — | — | — | 8.77(4.95) |
| *Random Effects* | Variance | Variance | Variance | Variance |
| Intercept | 14.24 | 14.34 | 12.21 | 11.05 |
| Residual | 10.05 | 9.45 | 7.10 | 7.10 |
| Time (Year) | — | — | 0.67 | 0.66 |
| *Goodness of Fit* |  |  |  |  |
| AIC | 3582.6 | 3550.3 | 3494.1 | 3484.7 |
| BIC | 3596.1 | 3568.2 | 3520.9 | 3529.4 |
| Deviance | 3576.6 | 3542.3 | 3482.1 | 3464.7 |
| Log Likelihood | -1788.3 | -1771.1 | -1741 | -1732.3 |
| Parameters | 3 | 4 | 6 | 10 |

Note. \*p < .05, \*\*p < .01, \*\*\*p < .001.

## Discussion

This study examined spatio-temporal patterns of EDOOD visits across Virginia, as well as socio-ecological factors associated with these patterns using techniques from statistics, data mining, and geographical information systems (GIS). Our spatial analysis revealed that EDOOD visit rates significantly varied across Virginia counties with clusters of overdose hotspots (primarily southwestern region) and coldspots (primarily northern region). Clustering analysis helped identify 5 distinct groups of counties based on the magnitude and the direction of change of the EDOOD visit rates over time. Although the overall trend of EDOOD visit rates differed in these groups, we observed rising trends in recent years (starting around 2019) and a slight decline in the visit rates from 2017 to 2018, in all the groups. The steepest rise in the EDOOD visit rates was seen in counties belonging to Group C (e.g., Grayson & Galax, Martinsville & Henry). Finally, the multilevel analysis revealed that the changes in the EDOOD visit rates were significantly associated with the changes in clinical care factors (i.e., access to care and quality of care) and socio-economic factors (i.e., levels of education, employment, income, family and social support, and community safety).

As aforementioned, hotspots of EDOOD visit rates were the most prominent and consistent in the southwestern part of Virginia. Southwest Virginia is a rural area where residents have higher morbidity and mortality rates often correlated to a shortage of healthcare services [49]. Residents in southwest Virginia often cannot afford annual health insurance deductibles and many medical expenses are not covered by insurance [50]. A portion of individuals reported only seeking healthcare as a last resort and many did not receive regular care from a health provider. Individuals in southwestern counties with lower access to care and quality of care, therefore, may be more at risk of opioid overdose. For example, Martinsville County had one of the highest EDOOD rates throughout the six-year period. On the other hand, many counties in northern Virginia had low EDOOD visit rates throughout the 6-year period. This region includes counties that are close to the capital and is considered to have good access to healthcare, better employment opportunities, and higher levels of education [38].

Study findings also revealed sub-groups of counties with similar EDOOD visit trends. Spatial mapping of these counties in Fig 3 indicates that many counties that belong to the same group are clustered together in space suggesting a possible association with the neighborhood characteristics. In most counties, the EDOOD visit rates decreased from 2017 to 2018. This is consistent with the national pattern and is believed to be attributed to the reduced prescribing volume of high-dose opioid pills and a sudden decline in the availability of a highly potent synthetic opioid (carfentanil) [20]. The increase in EDOOD visits from 2019 may depict how Covid-19 influenced the overall opioid overdose trends. Research conducted across six health care systems in the US [51] identified that EDOOD visit counts increased by 10.5% in 2020 compared to the counts in 2018 and 2019 despite having a 14% decrease in the overall ED visits. The same study pointed out that this rise might be attributed to the disruption of access to treatment, social support, loss of employment, social isolation, and many more during the Covid-19 period. Further, this increase in EDOOD rates from 2019 may reflect the increased prevalence and use of illicitly manufactured fentanyl, which has been the major driver of the opioid epidemic over the past few years (having surpassed prescription opioids and heroin as major causes of opioid overdose) [52]. This may also explain the sharp, increasing rates of EDOOD visits (throughout the 6-year period) in some Virginia counties (Group C in Fig 3).

Finally, our multilevel analysis found that a decrease in socio-economic factors over time was associated with increased EDOOD visit rates. This finding is consistent with studies that demonstrate associations between poor economic and social conditions and high opioid overdose mortality rates [5-6]. Conversely, the analysis revealed that counties with poor access to care and quality of care (i.e., higher clinical care z-scores) had lower EDOOD visit rates. It is possible that the inaccessibility and poor quality of clinical care might have led to individuals not being able to seek care or go to the ED. These findings suggest that addressing county-level deficits in clinical care and socioeconomic factors may help reduce opioid overdoses.

Some limitations of this study could be addressed in the future. If the necessary data is available, the study can be expanded to include a broader time frame and nationwide data. Additionally, breaking down the EDOOD visits by the type of opioids could provide a better picture of the epidemic which was not possible with the data that we used. Lastly, the spatio-temporal variations of EDOOD visits were assessed separately but they could be modeled together to identify how these interact with other outcomes and with each other.

## Conclusion

Overall, there are differences between the counties in Virginia in their EDOOD visit patterns across time. These differences are significantly associated with socio-economic factors (i.e., education, employment, community safety, income, and family and social support) and clinical care (i.e., access to care and quality of care). Targeting areas that are consistently hot spots for EDOOD rates and identifying areas that vary over time is critical to address the social determinants of opioid use disorders and health care access.

## Data Availability

The information about all relevant data are within the manuscript

## Acknowledgments

We would like to thank the faculty members and students who are a part of the NSF Research Traineeship (NRT) program at George Mason University for providing us with a collaborative environment, and for all the help and guidance provided to conduct this interdisciplinary research project. We would also like to extend our thanks to Sulabh Shrestha (GMU, Computer Science) for proofreading the manuscript.

## Declarations

### Data availability statement

The data used for this study are publicly available and are referenced in the paper.

## Appendix Virginia localities combined for EDOOD visit rate calculation

- Alleghany County and Covington City
- Albemarle County and Charlottesville City
- Augusta County, Staunton City, and Waynesboro City
- Chesterfield County and Colonial Heights City
- Frederick County and Winchester City
- Fairfax County, Fairfax City, and Falls Church City
- Grayson County and Galax City
- Greensville County and Emporia City
- Henry County and Martinsville City
- Montgomery County and Radford City
- Pittsylvania County and Danville City
- Prince George County, Hopewell City, and Petersburg City
- Prince William County, Manassas City, and Manassas Park City
- Roanoke County, Roanoke City, and Salem City
- Rockingham County and Harrisonburg City
- Rockbridge County, Buena Vista City, and Lexington City
- Southampton County and Franklin City
- Washington County and Bristol City
- Wise County and Norton City

## References

1. Seltzer N. (2020). The economic underpinnings of the drug epidemic. SSM - population health, 12, 100679. https://doi.org/10.1016/j.ssmph.2020.100679

2. Florence, C., Luo, F., & Rice, K. (2021). The economic burden of opioid use disorder and fatal opioid overdose in the United States, 2017. Drug and Alcohol Dependence, 218, 108350. https://doi.org/10.1016/j.drugalcdep.2020.108350

3. Centers for Disease Control and Prevention (2021). Drug Overdose Deaths in the U.S. Top 100,000 Annually. https://www.cdc.gov/nchs/pressroom/nchs_press_releases/2021/20211117.htm

4. Tracking federal funding to combat the opioid crisis. Safe States Resources Hub. (2022, May 12). https://resources.safestates.org/publications/tracking-federal-funding-to-combat-the-opioid-crisis

5. Altekruse, S. F., Cosgrove, C. M., Altekruse, W. C., Jenkins, R. A., & Blanco, C. (2020). Socioeconomic risk factors for fatal opioid overdoses in the United States: Findings from the Mortality Disparities in American Communities Study (MDAC). PloS One, 15(1), e0227966. https://doi.org/10.1371/journal.pone.0227966

6. Langabeer, J. R., Chambers, K. A., Cardenas-Turanzas, M., & Champagne-Langabeer, T. (2020). County-level factors underlying opioid mortality in the United States. Substance Abuse, 1–7. https://doi.org/10.1080/08897077.2020.1740379

7. Kamimura, A., Panahi, S., Rathi, N., Weaver, S., Pye, M., Sin, K., & Ashby, J. (2020). Risks of opioid abuse among uninsured primary care patients utilizing a free clinic. Journal of Ethnicity in Substance Abuse, 19(1), 58–69. https://doi.org/10.1080/15332640.2018.1456387

8. Linas, B. S., Latkin, C., Genz, A., Westergaard, R. P., Chang, L. W., Bollinger, R. C., & Kirk, G. D. (2015). Utilizing mHealth methods to identify patterns of high risk illicit drug use. Drug and Alcohol Dependence, 151, 250–257. https://doi.org/10.1016/j.drugalcdep.2015.03.031

9. Bohnert, A. S. B., Valenstein, M., Bair, M. J., Ganoczy, D., McCarthy, J. F., Ilgen, M. A., & Blow, F. C. (2011). Association between opioid prescribing patterns and opioid overdose-related deaths. JAMA: The Journal of the American Medical Association, 305(13), 1315–1321. https://doi.org/10.1001/jama.2011.370

10. Hall, A. J., Logan, J. E., Toblin, R. L., Kaplan, J. A., Kraner, J. C., Bixler, D., Crosby, A. E., & Paulozzi, L. J. (2008). Patterns of abuse among unintentional pharmaceutical overdose fatalities. JAMA: The Journal of the American Medical Association, 300(22), 2613–2620. https://doi.org/10.1001/jama.2008.802

11. Keyes, K. M., Cerdá, M., Brady, J. E., Havens, J. R., & Galea, S. (2014). Understanding the rural-urban differences in nonmedical prescription opioid use and abuse in the United States. American Journal of Public Health, 104(2), e52–e59. https://doi.org/10.2105/AJPH.2013.301709

12. Roehler, D. R., Guy, G. P., Jr, & Jones, C. M. (2020). Buprenorphine prescription dispensing rates and characteristics following federal changes in prescribing policy, 2017-2018: A cross-sectional study. Drug and Alcohol Dependence, 213, 108083. https://doi.org/10.1016/j.drugalcdep.2020.108083

13. Jalali, M. S., Botticelli, M., Hwang, R. C., Koh, H. K., & McHugh, R. K. (2020). The opioid crisis: A contextual, social-ecological framework. Health Research Policy and Systems, 18(1), 87. https://doi.org/10.1186/s12961-020-00596-8

14. Haffajee, R. L., Lin, L. A., Bohnert, A. S. B., & Goldstick, J. E. (2019). Characteristics of US Counties With High Opioid Overdose Mortality and Low Capacity to Deliver Medications for Opioid Use Disorder. JAMA Network Open, 2(6), e196373. https://doi.org/10.1001/jamanetworkopen.2019.6373

15. Monnat, S. M., Peters, D. J., Berg, M. T., & Hochstetler, A. (2019). Using census data to understand county-level differences in overall drug mortality and opioid-related mortality by opioid type. American Journal of Public Health, 109(8), 1084–1091. https://doi.org/10.2105/AJPH.2019.305136

16. James, C. V., Moonesinghe, R., Wilson-Frederick, S. M., Hall, J. E., Penman-Aguilar, A., & Bouye. (2017). Racial/ethnic health disparities among rural adults—United States, 2012–2015. MMWR. Surveillance Summaries, 66(23), 1–99. https://doi.org/10.15585/mmwr.ss6623a1

17. Marotta, P. L., Hunt, T., Gilbert, L., Wu, E., Goddard-Eckrich, D., & El-Bassel, N. (2019). Assessing spatial relationships between prescription drugs, race, and overdose in New York State from 2013 to 2015. Journal of Psychoactive Drugs, 51(4), 360–370. https://doi.org/10.1080/02791072.2019.1599472

18. Maus, M. (2015). Reconnecting public health and urban planning: An exploratory study of cross-agency collaboration. UC Berkeley, 75(8-B(E)).

19. Post, L. A., Lundberg, A., Moss, C. B., Brandt, C. A., Quan, I., Han, L., & Mason, M. (2022). Geographic Trends in Opioid Overdoses in the US From 1999 to 2020. JAMA Network Open, 5(7), e2223631. https://doi.org/10.1001/jamanetworkopen.2022.23631

20. Ciccarone, D. (2021). The rise of illicit fentanyls, stimulants and the fourth wave of the opioid overdose crisis. Current Opinion in Psychiatry, 34(4), 344–350. https://doi.org/10.1097/YCO.0000000000000717

21. Hernandez, A., Branscum, A. J., Li, J., MacKinnon, N. J., Hincapie, A. L., & Cuadros, D. F. (2020). Epidemiological and geospatial profile of the prescription opioid crisis in Ohio, United States. Scientific Reports, 10(1), 4341. https://doi.org/10.1038/s41598-020-61281-y

22. Sauer, J., Stewart, K., & Dezman, Z. D. W. (2021). A spatio-temporal Bayesian model to estimate risk and evaluate factors related to drug-involved emergency department visits in the greater Baltimore metropolitan area. Journal of Substance Abuse Treatment, 131, 108534. https://doi.org/10.1016/j.jsat.2021.108534

23. Park, C., Clemenceau, J. R., Seballos, A., Crawford, S., Lopez, R., Coy, T., Atluri, G., & Hwang, T. H. (2021). A spatiotemporal analysis of opioid poisoning mortality in Ohio from 2010 to 2016. Scientific Reports, 11(1), 4692. https://doi.org/10.1038/s41598-021-83544-y

24. Cao, Y., Stewart, K., Factor, J., Billing, A., Massey, E., Artigiani, E., Wagner, M., Dezman, Z., & Wish, E. (2020). Using socially-sensed data to infer ZIP level characteristics for the spatiotemporal analysis of drug-related health problems in Maryland. Health & Place, 63, 102345. https://doi.org/10.1016/j.healthplace.2020.102345

25. Kline, D., Pan, Y., & Hepler, S. A. (2021). Spatiotemporal Trends in Opioid Overdose Deaths by Race for Counties in Ohio. Epidemiology, 32(2), 295–302. https://doi.org/10.1097/EDE.0000000000001299

26. [dataset] Virginia Department of Health. (2021a). Fatal drug overdose quarterly report 2nd quarter 2021. Forensic Epidemiology. https://www.vdh.virginia.gov/content/uploads/sites/18/2021/10/Quarterly-Drug-Death-Report-FINAL-Q2-2021-1.pdf

27. Nolan, M. L., Kunins, H. V., Lall, R., & Paone, D. (2017). Developing syndromic surveillance to monitor and respond to adverse health events related to psychoactive substance use. Public Health Reports (1974-), 132(1S), 65S–72S. JSTOR. https://doi.org/10.2307/26374204

28. Hawk, K., & D’Onofrio, G. (2018). Emergency department screening and interventions for substance use disorders. Addiction Science & Clinical Practice, 13(1), 18. https://doi.org/10.1186/s13722-018-0117-1

29. FAIR Health, Inc. (2017). Peeling back the curtain on regional variation in the opioid crisis: Spotlight on five key urban centers and their respective stated. https://collections.nlm.nih.gov/catalog/nlm:nlmuid-101751557-pdf

30. Mayo Clinic. (2021). Emergency department visits related to opioid overdoses up significantly during COVID-19 pandemic. ScienceDaily. Retrieved from http://www.sciencedaily.com/releases/2021/07/210728201358.htm

31. Zhou, X., & Lin, H. (2008). Morans I. In S. Shekhar & H. Xiong (Eds.), Encyclopedia of GIS (pp. 725–725). Springer US. https://doi.org/10.1007/978-0-387-35973-1_817

32. Anselin, L. (1995). Local indicators of spatial association—LISA. Geographical Analysis, 27(2), 93–115. https://doi.org/10.1111/j.1538-4632.1995.tb00338.x

33. Müller, M. (2007). Dynamic time warping. In Information Retrieval for Music and Motion (pp. 69–84). Springer-Verlag. https://doi.org/10.1007/978-3-540-74048-3_4

34. Mumper, M. (2017). Multilevel modelling. American Psychological Association. https://www.apa.org/science/about/psa/2017/01/multilevel-modelling

35. [dataset] Virginia Department of Health. (2021b). Monthly and annual statistics, 2015 – 2021. ED Visits for Drug Overdose. https://www.vdh.virginia.gov/surveillance-and-investigation/syndromic-surveillance/drug-overose-surveillance/

36. Drug overdose case definitions. Surveillance and Investigation. (2022, June 16). Retrieved September 18, 2022, from https://www.vdh.virginia.gov/surveillance-and-investigation/syndromic-surveillance/drug-overdose-case-definition/

37. ACS public use microdata sample (PUMS) overview. Washington, D.C. :U.S. Census Bureau.

38. [dataset] Kingery, K. L. (2018). County health rankings & roadmaps. Journal of Youth Development, 13(3), 259–263. https://doi.org/10.5195/jyd.2018.649

39. [dataset] County Health Rankings. (2020). County health rankings and roadmaps. County Health Rankings & Roadmaps. https://www.countyhealthrankings.org/county-health-rankings-roadmaps

40. Anderson, T. J., Saman, D. M., Lipsky, M. S., & Lutfiyya, M. N. (2015). A cross-sectional study on health differences between rural and non-rural U.S. counties using the County Health Rankings. BMC Health Services Research, 15(1), 441. https://doi.org/10.1186/s12913-015-1053-3

41. Remington, P. L., Catlin, B. B., & Gennuso, K. P. (2015). The County Health Rankings: Rationale and methods. Population Health Metrics, 13(1), 11. https://doi.org/10.1186/s12963-015-0044-2

42. [dataset] U.S. Census Bureau. (2018). County adjacency file. United States Census Bureau. https://www.census.gov/geographies/reference-files/2010/geo/county-adjacency.html

43. Getis, A. (1995). Cliff, A.D. and Ord, J.K. 1973: Spatial autocorrelation. London: Pion. Progress in Human Geography, 19(2), 245–249. https://doi.org/10.1177/030913259501900205

44. Rey, S. J., & Anselin, L. (2007). PySAL: A Python library of spatial analytical methods. Review of Regional Studies, 37(1), 5–27. https://doi.org/10.52324/001c.8285

45. Tavenard, R., Faouzi, J., Vandewiele, G., Divo, F., Androz, G., Holtz, C., Payne, M., Yurchak, R., Rußwurm, M., Kolar, K., & Woods, E. (2020). Tslearn, a machine learning toolkit for time series data. Journal of Machine Learning Research, 21(118), 1–6.

46. Bates, D., Mächler, M., Bolker, B., & Walker, S. (2015). Fitting linear mixed-effects models using lme4. Journal of Statistical Software, 67, 1–48. https://doi.org/10.18637/jss.v067.i01

47. Nezlek, J. B. (2008). An introduction to multilevel modeling for social and personality psychology. Social and Personality Psychology Compass, 2(2), 842–860. https://doi.org/10.1111/j.1751-9004.2007.00059.x

48. Bryk, A. S., & Raudenbush, S. W. (1992). Hierarchical linear models: Applications and data analysis methods. Sage Publications, Inc.

49. Merwin, E., Snyder, A., & Katz, E. (2006). Differential access to quality rural healthcare: Professional and policy challenges. Family & Community Health, 29(3), 186–194. https://doi.org/10.1097/00003727-200607000-00005

50. Huttlinger, K., Schaller-Ayers, J., & Lawson, T. (2004). Health care in Appalachia: A population-based approach. Public Health Nursing, 21(2), 103–110. https://doi.org/10.1111/j.0737-1209.2004.021203.x

51. Soares, W. E., Melnick, E. R., Nath, B., DOnofrio, G., Paek, H., Skains, R. M., Walter, L. A., Casey, M. F., Napoli, A., Hoppe, J. A., & Jeffery, M. M. (2021). Emergency Department Visits for Nonfatal Opioid Overdose During the COVID-19 Pandemic Across Six US Health Care Systems. Annals of Emergency Medicine, S0196064421002262. https://doi.org/10.1016/j.annemergmed.2021.03.013

52. Hedegaard H, Miniño AM, Spencer MR, Warner M. Drug overdose deaths in the United States, 1999–2020. NCHS Data Brief, no 428. Hyattsville, MD: National Center for Health Statistics. 2021. DOI: https://dx.doi.org/10.15620/cdc:112340

